# Do we need to reconsider knee valgus as a sole risk factor for second ACL injury? Exploratory analysis of individual Landing Error Scoring System (LESS) items with three year follow-up

**DOI:** 10.1101/2023.07.31.23293477

**Authors:** Nicky van Melick, Lisa van Rijn, Rob Bogie, Thomas J. Hoogeboom

## Abstract

**Background:** It remains unclear which movement quality items are associated with second anterior cruciate ligament (ACL) injuries after ACL reconstruction.

**Hypotheses:** 1) Movement quality measured with the Landing Error Scoring System (LESS) is not associated with second ACL injury in soccer players who returned to sports after ACL reconstruction. 2) In an exploratory analysis, soccer players with and without second ACL injuries display differences in individual LESS items.

**Methods:** Fourteen male recreational ACL-reconstructed soccer players had non-fatigued and fatigued movement quality measurement with LESS score three years ago and were now interviewed by phone about soccer participation and second ACL injuries.

**Results:** Two soccer players did not return to soccer and were therefore excluded from analysis. Six of twelve (50%) included soccer players had a second ACL injury. There was no association between the non-fatigued (p=1.000) and fatigued (p=0.455) LESS score and second ACL injuries. However, when exploring individual LESS items, we found 50% of the soccer players with second ACL injury landing with little trunk and knee flexion displacement in combination with knee valgus, while none of the soccer players without second ACL injury had this combination of LESS errors. This difference was only seen in the fatigued state.

**Conclusions:** It might be the combination of little knee and trunk flexion displacement with knee valgus during landing which predisposes an athlete to a second ACL injury, especially in a fatigued state.

**Clinical relevance:** When screening athletes after ACL reconstruction for second ACL injury risk, combining measurement of knee valgus with knee and trunk flexion displacement in a fatigued state might be give more insight into risk than using the complete LESS score or evaluating knee valgus as a sole risk factor.

## 1. Introduction

Movement quality has become an increasingly popular tool to guide return to sport (RTS) decisions for athletes who want to return to pivoting sports after anterior cruciate ligament reconstruction (ACLR).^2,13^ Several options are available to measure movement quality, ranging from lab-based measurements to relatively easy-to-use on-field measurements. An example of a frequently used movement quality measurement is the Landing Error Scoring System (LESS) which consists of 17 movement quality items, with a lower score representing better movement quality.^9^ Padua et al. found an association between LESS score and primary ACL injuries in adolescent soccer athletes, and suggested a score of 5 as the optimal cut-off point, with a sensitivity of 85% and a specificity of 64%.^10^ We, however, found no association between LESS score and second ACL injuries in adult athletes returning to pivoting sports after ACLR.^15^ Despite the lack of scientific support for the predictive value of the LESS for second ACL injuries, many clinicians use the LESS as an instrument to determine whether an athlete is ready to safely return to pivoting sports after ACLR anyway.

Perhaps, to maximize the predictive value of the LESS we need to consider other ways of analysing or applying the LESS. Two new approaches come to mind.

First, individual LESS items or a combination of a few items might demonstrate a stronger predictive value for second ACL injuries than all LESS items together. Knee valgus is one of the items that clinicians always mention as being important in preventing second ACL injuries, but the only scientific evidence comes from studies focussing on primary ACL injuries.^8^ Studies looking at second ACL injuries only showed that less hip external rotator moment during early landing and little knee flexion displacement during hop-and-hold test are important associated items.^11,15^

Second, application of the LESS in fatigued athletes might also increase its predictive capacities. In a recent cross-sectional study we found LESS score measured on the soccer field to increase more in ACL-reconstructed recreational soccer players than in healthy recreational soccer players when comparing a sport-specific fatigued state to a non-fatigued state. We hypothesized that a higher fatigued LESS score might predispose the ACL-reconstructed soccer players to a second ACL injury.^14^

In this exploratory study we aimed to investigate the plausibility of these new approaches of analyzing and applying the LESS. The first goal was to assess whether movement quality (measured on the soccer field with LESS score) in a non-fatigued or fatigued state is associated with second ACL injuries during a three-year follow-up in ACL-reconstructed male recreational soccer players who returned to soccer. The second goal was to explore whether individual LESS items or a combination of a few items - in both the non-fatigued and fatigued state – differed between soccer players who had a second ACL injury compared to those who did not have second ACL injury.

## 2. Materials and Methods

This retrospective exploratory case series is a three-year follow-up study of a previously published cross-sectional study comparing the effects of sport-specific fatigue on movement quantity and quality RTS measurements in healthy and ACL-reconstructed recreational soccer players.^14^ RTS measurements were all performed on the soccer field and were taken at the end of rehabilitation (12.4 ± 3.5 months after surgery) for all ACL-reconstructed soccer players. Apart from a double-leg countermovement jump with LESS score, RTS measurements also included a single-leg hop test battery: vertical jump, hop for distance, and side hop.^14^ The LESS has a good to excellent intrarater (ICC 0.82-0.99) and interrater (ICC 0.83-0.92) reliability.^6^ All RTS measurements were performed in a non-fatigued state and in a sport-specific fatigued state. Fatigue was induced by a one-hour soccer training and the Borg RPE scale was used to express the amount of fatigue. Borg RPE scale was 7.3 ± 1.1 during the non-fatigued RTS measurements and 14.9 ± 1.1 during the fatigued measurements.^14^

### 2.1 Participants and study procedure

Three years after RTS measurements we asked all 14 ACL-reconstructed soccer players by phone to participate in this follow-up study. They received additional information about the study by e-mail and decided whether they wanted to partake afterwards. Once a soccer player agreed to participate in this follow-up study, a 10-minute interview by phone was scheduled. A standardised survey about soccer participation and second ACL injuries was used to guide this interview.

All subjects gave informed consent for participation in this follow-up study. The medical ethics committee of the Máxima Medisch Centrum Eindhoven (the Netherlands) deemed that our study did not fall within the remit of the Medical Research Involving Human Subjects Act (N19.117).

### 2.2 Statistical analyses

There were no missing values. For all soccer players who returned to soccer, descriptive statistics were used to present age, height, weight, operated leg, dominant leg, soccer exposure in months, non-fatigued LESS score and fatigued LESS score.

To analyse differences in baseline characteristics for soccer players who had a second ACL injury and those who had not independent samples t-tests (continuous variables) or chi-square tests (nominal variables) were performed. Soccer players who did not return to soccer (or any other pivoting sport) were excluded from analyses, because they had a lower chance for a second ACL injury attributable to much lower risk exposure.

The association between non-fatigued or fatigued LESS score and second ACL injury rate when returning to soccer was investigated using Fisher’s exact test. LESS score was expressed as a dichotomous variable (meeting RTS criterion of ≤5 yes or no). Analysis were performed for the non-fatigued and fatigued state separately. The absolute risk (AR) was calculated with crosstabs, comparing soccer players who did not meet the LESS criterion of ≤5 with those who did meet the LESS criterion.

Individual LESS items were analysed in an explorative way. For every item, both in the non-fatigued and fatigued state, the number and percentage of soccer players with and without second ACL injury not meeting the item was described. We arbitrarily deemed a ≥50% difference between groups sufficient to identify clinically meaningful differences and to minimise findings based on chance.

## 3. Results

All 14 ACL-reconstructed soccer players agreed to participate in this follow-up study. Two of them had not returned to soccer (or any other pivoting sport) after ACLR due to complaints of the operated knee and were therefore excluded from analyses. The other 12 ACL-reconstructed soccer players returned to recreational level competitive soccer and were included in the analyses (Figure 1).

**Figure 1:**
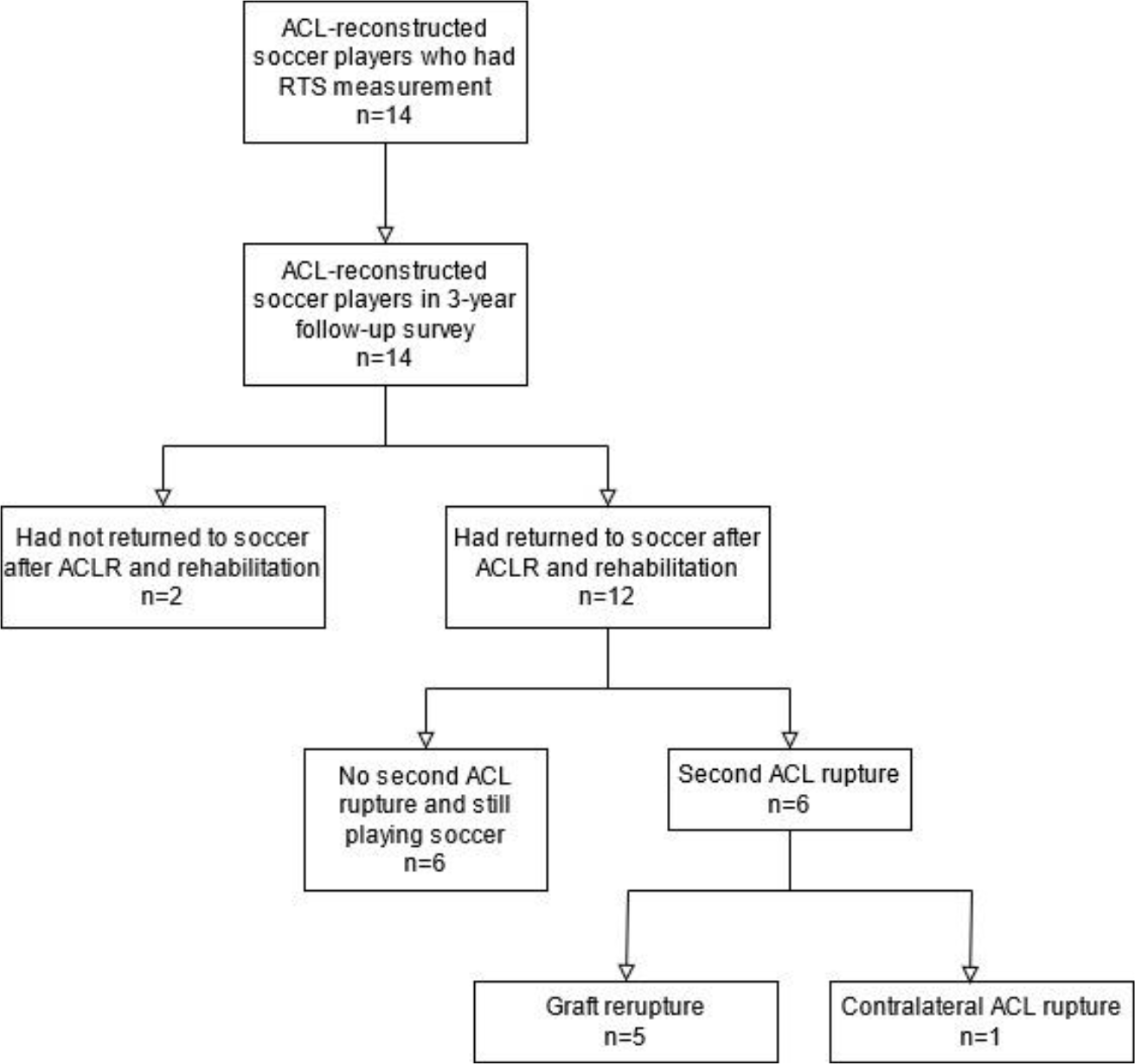
Flowchart of ACL-reconstructed soccer players.

### 3.1 Second ACL injuries

Descriptives for all ACL-reconstructed soccer players separately are presented in Table 1. Six soccer players (50%) had a second ACL injury during the three-year follow-up period. Five were non-contact graft reruptures and one was a non-contact contralateral ACL rupture (soccer player 6). Second ACL ruptures occurred between 1 and 24 months after returning to soccer (median 4.5 months). Four soccer players ruptured their ACL during training, the other two during a game. Second ACL injuries happened within 30 to 80 minutes (median 60 minutes) from the beginning of training or game.

**Table 1:** Descriptives for all ACL-Reconstructed Soccer Players separately.

| Soccer player | Second ACL rupture? | Soccer exposure in months | Reinjury during game or training? | In-game/training timing in minutes | Non-fatigued LESS score | Fatigued LESS score |
| --- | --- | --- | --- | --- | --- | --- |
| 1 | Yes | 1 | Training | 80 | 3 | 3 |
| 2 | Yes | 24 | Training | 30 | 6 | 10 |
| 3 | Yes | 2 | Game | 70 | 1 | 6 |
| 4 | Yes | 8 | Game | 70 | 4 | 7 |
| 5 | Yes | 6 | Training | 50 | 4 | 8 |
| 6 | Yes | 3 | Training | 40 | 4 | 6 |
| 7 | No | 34 | n.a. | n.a. | 4 | 6 |
| 8 | No | 35 | n.a. | n.a. | 2 | 6 |
| 9 | No | 36 | n.a. | n.a. | 4 | 7 |
| 10 | No | 36 | n.a. | n.a. | 4 | 6 |
| 11 | No | 34 | n.a. | n.a. | 2 | 6 |
| 12 | No | 36 | n.a. | n.a. | 5 | 8 |

### 3.2 Association between LESS score and second ACL injuries

Characteristics of soccer players with second ACL injury did not significantly differ from soccer players without second ACL injury (Table 2). In the non-fatigued state one soccer player did not meet the LESS criterion of ≤5 and he had a second ACL injury. In the fatigued state one soccer players did meet the LESS RTS criterion, but he had a second ACL injury (Table 1). Fisher’s exact test showed no association between not meeting the non-fatigued (p=1.00; AR 100% versus 45%) or fatigued (p=0.46; AR 45% versus 100%) LESS RTS criterion and second ACL injuries.

**Table 2:** Characteristics of ACL-Reconstructed Soccer Players with and without Second ACL Injury.

|  | Soccer players with second ACL injury | Soccer players without second ACL injury | p-value |
| --- | --- | --- | --- |
| Age in years, mean ± SD | 20.7 ± 2.7 | 23.8 ± 2.9 | 0.130 |
| Height in m, mean ± SD | 181.3 ± 3.2 | 177.8 ± 8.7 | 0.378 |
| Weight in kg, mean ± SD | 80.7 ± 5.3 | 75.5 ± 10.5 | 0.308 |
| ACLR leg, N (%) right | 2 (33%) | 3 (50%) | 0.558 |
| Dominant leg = operated leg?, N (%) yes | 2 (33%) | 4 (66%) | 0.248 |
| Non-fatigued LESS score, mean ± SD | 3.7 ± 1.6 | 3.5 ± 1.2 | 0.845 |
| Fatigued LESS score, mean ± SD | 6.7 ± 2.3 | 6.5 ± 0.8 | 0.873 |

### 3.3 Exploratory analysis of LESS items

Exploring individual LESS items, three ≥50% differences were found between soccer players with second ACL injury and those without second ACL injury, all three in the fatigued state (Table 3).

**Table 3:** Exploratory Analysis of Individual LESS Items and Differences between Soccer Players with and without Second ACL Injury.

| State | LESS item | Not meeting item if | Number (%) of soccer players with second ACL injury not meeting item | Number (%) of soccer players without second ACL injury not meeting item |
| --- | --- | --- | --- | --- |
| Non-fatigued | 1 | Knee flexion angle $\leq 30^\circ$ (initial contact) | 0 (0) | 1 (17) |
|  | 2 | The thigh of test leg is in line with the trunk (initial contact) | 0 (0) | 0 (0) |
|  | 3 | The trunk is vertical or extended on the hips (initial contact) | 0 (0) | 0 (0) |
|  | 4 | If the foot lands heel to toe or flat | 0 (0) | 0 (0) |
|  | 5 | A line straight down from the center of the patella is medial to the midfoot (initial contact) | 2 (33) | 3 (50) |
|  | 6 | The midline of the trunk is flexed to the right or left side (initial contact) | 4 (67) | 2 (33) |
|  | 7 | A line down from the tip of the shoulders is inside the foot (entire foot contact) | 3 (50) | 3 (50) |
|  | 8 | A line down from the tip of the shoulders is outside the foot (entire foot contact) | 2 (33) | 1 (17) |
| | 9 | The foot is internally rotated $>30^\circ$ (between initial contact and maximal knee flexion) | 0 (0) | 0 (0) |
| | 10 | The foot is externally rotated $>30^\circ$ (between initial contact and maximal knee flexion) | 0 (0) | 0 (0) |
|  | 11 | One foot lands before the other or if one foot lands heel to toe and the other toe to heel | 1 (17) | 2 (33) |
| | 12 | The knee flexes $<45^\circ$ (between initial contact and maximal knee flexion) | 2 (33) | 0 (0) |
|  | 13 | The thigh does not flex more on the trunk (between initial contact and maximal knee flexion) | 0 (0) | 0 (0) |
|  | 14 | The trunk does not flex more (between initial contact and maximal knee flexion) | 0 (0) | 0 (0) |
|  | 15 | A line straight down from the center of the patella runs through or is medial to the great toe (at maximum knee valgus) | 5 (83) | 4 (67) |
| Fatigued | 1 | Knee flexion angle $\leq 30^\circ$ (initial contact) | 1 (17) | 4 (67) |
|  | 2 | The thigh of test leg is in line with the trunk (initial contact) | 0 (0) | 1 (17) |
|  | 3 | The trunk is vertical or extended on the hips (initial contact) | 1 (17) | 1 (17) |
|  | 4 | If the foot lands heel to toe or flat | 0 (0) | 1 (17) |
|  | 5 | A line straight down from the center of the patella is medial to the midfoot (initial contact) | 4 (67) | 3 (50) |
|  | 6 | The midline of the trunk is flexed to the right or left side (initial contact) | 3 (50) | 3 (50) |
|  | 7 | A line down from the tip of the shoulders is inside the foot (entire foot contact) | 2 (33) | 4 (67) |
|  | 8 | A line down from the tip of the shoulders is outside the foot (entire foot contact) | 3 (50) | 1 (17) |
| | 9 | The foot is internally rotated $>30^\circ$ (between initial contact and maximal knee flexion) | 0 (0) | 0 (0) |
| | 10 | The foot is externally rotated $>30^\circ$ (between initial contact and maximal knee flexion) | 2 (33) | 2 (33) |
|  | 11 | One foot lands before the other or if one foot lands heel to toe and the other toe to heel | 1 (17) | 3 (50) |
| | 12 | The knee flexes $<45^\circ$ (between initial contact and maximal knee flexion) | 4 (67) | 0 (0) |
|  | 13 | The thigh does not flex more on the trunk (between initial contact and maximal knee flexion) | 1 (33) | 0 (0) |
|  | 14 | The trunk does not flex more (between initial contact and maximal knee flexion) | 3 (50) | 0 (0) |
|  | 15 | A line straight down from the center of the patella runs through or is medial to the great toe (at maximum knee valgus) | 5 (83) | 5 (83) |

In the fatigued state soccer players with second ACL injury:

- less often had a small knee flexion angle (≤30°) at initial contact compared to those without second ACL injury (Item 1: 17% respectively 67%);
- more often had little knee flexion displacement (<45°) from initial contact to maximal knee flexion compared to those without second ACL injury (Item 12: 67% respectively 0%);
- more often had no trunk flexion displacement between initial contact and maximal knee flexion compared to those without second ACL injury (Item 14: 50% respectively 0%).

The occurrence of knee valgus (both item 5 and 15) did not differ between soccer players with or without second ACL injury, nor in the non-fatigued and in the fatigued state.

When combining items, three (50%) of the soccer players with second ACL injury had an error on item 12, 14 and 15, indicating little knee and trunk flexion displacement combined with knee valgus, while non of the soccer players without second ACL injury had this combination of items (see Appendix).

## 4. Discussion

This study showed there is no association between non-fatigued and fatigued LESS score and second ACL injuries in ACL-reconstructed recreational soccer players who returned to soccer. However, when exploring individual LESS items or a combination of a few items in the fatigued state, 50% of the soccer players with second ACL injury had little knee and trunk flexion displacement combined with knee valgus during landing. None of the soccer players without second ACL injury had this combination of items: they also had knee valgus, but with greater knee and trunk flexion displacement. Interestingly, soccer players with second ACL injury more often had a greater knee flexion angle at initial contact.

Despite the small sample of this exploratory cohort study, these findings are in line with a previous study of our group in which we found no association between LESS score and second ACL injury rate in adult athletes returning to pivoting sports after ACLR.^15^ In the same study, we found less knee flexion displacement during hop-and-hold test to be associated with second ACL injuries.^15^ Also, biomechanical studies have indicated that proximal factors influence knee loading, and that actively flexing the trunk produces concomitant increases in hip and knee flexion.^1,12^ Coupling of proximal and distal joints increases the demands on the hip extensors (i.e. gluteal muscles, hamstrings) and decreases the demands on the knee extensors and thereby lowers the strain on the ACL.^12^ ACL injuries happen short (40-70 ms) after initial contact and quick absorption of ground reaction forces by flexing the trunk and knee could potentially protect the ACL from rupturing, even when knee valgus is occurring.^1,3^ Probably, knee and trunk flexion at initial contact are of less importance, since more soccer players without second ACL injury had a small knee flexion angle at that time point compared to the soccer players with second ACL injury.

Since almost all soccer players in this study (83%) had knee valgus during landing in the fatigued state, this individual item might not be a sole risk factor for second ACL injury. It might be the combination of little knee and trunk flexion displacement and knee valgus during landing which predisposes an athlete to second ACL injury, especially in a fatigued state.

It could be questioned whether our group of ACL-reconstructed soccer players had low-quality rehabilitation. To be included in our previous study measuring RTS tests on the soccer field, soccer players were recruited via experienced sports physical therapists who worked according to ACLR practice guidelines.^13,14^ While quality of the rehabilitation process could never be completely transparent, we think these soccer players all had a high-quality rehabilitation, including strength training, neuromuscular training, on-field training and multiple test moments. However, in future more attention should be paid to avoiding an erect landing posture in combination with knee valgus, especially when being fatigued.

Besides, we postulate that well-known RTS test batteries including strength tests, hop tests, and movement quality measurements with LESS need to be critically reviewed. Since strength and hop tests have an association with second ACL injuries in athletes who return to pivoting sports in multiple studies, we believe these are minimal RTS requirements.^4,5,7,15^ When using movement quality RTS tests, we advise analyzing individual items in a (sport specific) fatigued state, because these individual LESS items might give more insight into second ACL injury risk.

## 5. Conclusion

There is no associated between non-fatigued or fatigued LESS score and second ACL injury in ACL-reconstructed recreational soccer players who returned to soccer. However, soccer players with second ACL injury more often had little knee and trunk flexion displacement in the fatigued state, while there were no differences in the occurrence of knee valgus.We postulate that knee valgus is not the sole factor in second ACL injury risk. It might be the combination of little knee and trunk flexion displacement with knee valgus during landing which predisposes an athlete to a second ACL injury, especially in a fatigued state.

## Data Availability

All data produced in the present work are contained in the manuscript.

---

**Appendix:** Exploratory analysis of individual LESS items per soccer player in the fatigued state.

|  | Soccer player |  |  |  |  |  |  |  |  |  |  |  |
| --- | --- | --- | --- | --- | --- | --- | --- | --- | --- | --- | --- | --- |
|  | 1 | 2 | 3 | 4 | 5 | 6 | 7 | 8 | 9 | 10 | 11 | 12 |
| LESS item 1 |  |  | X |  |  |  | X |  | X | X | X |  |
| LESS item 2 |  |  |  |  |  |  |  |  |  |  | X |  |
| LESS item 3 |  | X |  |  |  |  |  |  | X |  |  |  |
| LESS item 4 |  |  |  |  |  |  |  |  |  |  |  | X |
| LESS item 5 | X | X | X | X |  |  | X |  |  | X |  | X |
| LESS item 6 |  |  | X | X | X |  |  | X |  | X |  | X |
| LESS item 7 |  | X |  | X |  |  | X |  | X |  | X | X |
| LESS item 8 |  |  | X |  | X | X |  | X |  |  |  |  |
| LESS item 9 |  |  |  |  |  |  |  |  |  |  |  |  |
| LESS item 10 |  | X |  |  |  | X | X |  |  |  | X |  |
| LESS item 11 |  |  | X |  |  |  |  | X | X |  |  | X |
| LESS item 12 |  | X |  | X | X | X |  |  |  |  |  |  |
| LESS item 13 |  |  |  |  | X |  |  |  |  |  |  |  |
| LESS item 14 |  | X |  | X | X |  |  |  |  |  |  |  |
| LESS item 15 |  | X | X | X | X | X | X | X |  | X | X | X |

An "X" indicates an error on a specific LESS item.

## References

1. Blackburn JT, Padua DA. Influence of trunk flexion on hip and knee joint kinematics during a controlled drop landing. Clin Biomech. 2008;23(3):313–313.

2. Davies GJ, McCarty E, Provencher M, Manske RC. ACL Return to sport guidelines and criteria. Curr Rev Musculoskelet Med. 2017;10(3):307–314.

3. Della Villa F, Buckthorpe M, Grassi A, et al. Systematic video analysis of ACL injuries in professional male football (soccer): injury mechanisms, situational patterns and biomechanics study on 134 consecutive cases. Br J Sports Med. 2020;54(23):1423–1423.

4. Grindem H, Snyder-Mackler L, Moksnes H, Engebretsen L, Risberg MA. Simple decision rules can reduce reinjury risk by 84% after ACL reconstruction: the Delaware-Oslo ACL cohort study. Br J Sports Med. 2016;50(13):804–804.

5. Grindem H, Engebretsen L, Axe M, Snyder-Mackler L, Risberg MA. Activity and functional readiness, not age, are the critical factors for second anterior cruciate ligament injury - the Delaware-Oslo ACL cohort study. Br J Sports Med. 2020;54(18):1099–1099.

6. Hanzlíková I, Hébert-Losier K. Is the landing Error Scoring System reliable and valid? A systematic review. Sports Health. 2020;12(2):181–181.

7. Kyritsis P, Bahr R, Landreau P, Miladi R, Witvrouw E. Likelihood of ACL graft rupture: not meeting six clinical discharge criteria before return to sport is associated with a four times greater risk of rupture. Br J Sports Med. 2016;50(15):946–946.

8. Larwa J, Stoy C, Chafetz RS, Boniello M, Franklin C. Stiff landings, core stability, and dynamic knee valgus: a systematic review on documented anterior cruciate ligament ruptures in male and female athletes. Int J Environ Res Public Health. 2021;18(7):3826.

9. Padua DA, Marshall SW, Boling MC, Thigpen CA, Garrett Jr WE, Beutler AI. The Landing Error Scoring System (LESS) is a valid and reliable clinical assessment tool of jump-landing biomechanics: The JUMP-ACL study. Am J Sports Med. 2009;37(10):1996–1996.

10. Padua DA, DiStefano LJ, Beutler AI, de la Motte SJ, DiStefano MJ, Marshall SW. The Landing Error Scoring System as a screening tool for an anterior cruciate ligament injury-prevention program in elite youth soccer athletes. J Athl Train. 2015;50(6):589–589.

11. Paterno MV, Schmitt LC, Ford KR, et al. Biomechanical measures during landing and postural stability predict second anterior cruciate ligament injury after anterior cruciate ligament reconstruction and return to sport. Am J Sports Med. 2010;38(10):1968–1968.

12. Powers CM. The influence of abnormal hip mechanics on knee injury: a biomechanical perspective. J Orthop Sports Phys Ther. 2010;40(2):42–42.

13. van Melick N, van Cingel REH, Brooijmans F, et al. Evidence-based clinical practice update: practice guidelines for anterior cruciate ligament rehabilitation based on a systematic review and multidisciplinary consensus. Br J Sports Med. 2016;50(24):1506–1506.

14. van Melick N, van Rijn L, Nijhuis-van der Sanden MWG, Hoogeboom TH, van Cingel REH. Fatigue affects quality of movement more in ACL-reconstructed soccer players than in healthy soccer players. Knee Surg Sports Traumatol Arthrosc. 2019;27(2):549–549.

15. van Melick N, Pronk Y, Nijhuis-van der Sanden MWG, Rutten S, van Tienen TG, Hoogeboom TH. Meeting movement quantity or quality return to sport criteria is associated with reduced second ACL injury rate. J Orthop Res. 2022;40(1):117–117.

